# Visual Imagery and Spectrum Symptoms of Depression and Hypomania Differentially Modulate Brain Responses during Emotional Face Anticipation and Encoding

**DOI:** 10.1101/2025.11.05.25339588

**Authors:** Anna Manelis, Skye Satz, Rachel Miceli, Satish Iyengar, Holly A. Swartz

## Abstract

**Background:** Depressive (DD) and bipolar (BD) disorders are characterized by biases in anticipating and encoding emotional information. Depressive traits are linked to negative affective biases, while hypomanic features are associated with heightened responsiveness to positive stimuli. The vividness of visual imagery may further modulate these biases. This study examined how lifetime dimensional symptoms of depression and hypomania across diagnoses interact with imagery vividness to modulate brain activation during anticipation and encoding of happy and sad faces.

**Methods:** A total of 155 individuals aged 18–45 years with BD, DD, or healthy control (HC) status completed a cued emotional face-encoding task during functional magnetic resonance imaging (fMRI). Lifetime dimensional symptoms of depression and hypomania were assessed using the MOODS-SR, and imagery vividness was measured with the Vividness of Visual Imagery Questionnaire (VVIQ). Interaction effects between spectrum depression/hypomania and imagery vividness on brain activation during anticipation and encoding of happy versus sad faces were analyzed using the Sandwich Estimator approach in FSL.

**Results:** Higher spectrum hypomania scores were associated with greater imagery vividness and increased activation for happy versus sad faces in the occipital pole, cuneus, intracalcarine, and lateral occipital cortices during encoding. However, there was reduced activation for happy versus sad faces in the precentral gyrus during anticipation. Depressive symptoms interacted with imagery vividness within the default mode network: more severe spectrum depression and lower vividness were associated with greater activation for happy versus sad faces in the frontopolar cortex during anticipation, but with reduced activation in the angular gyrus during encoding.

**Conclusion:** Lifetime depressive and hypomanic spectrum symptoms across diagnoses differentially interact with visual imagery to influence anticipation and encoding of emotional faces. Depressive biases were observed in frontopolar–parietal regions, while hypomanic biases appeared in occipital cortices. These findings highlight the value of dimensional approaches to mood psychopathology and identify imagery vividness as a promising transdiagnostic target for intervention.

## 1 INTRODUCTION

Depressive (DD) and bipolar (BD) disorders are among the most disabling psychiatric conditions contributing to substantial personal and global health challenges [1–4]. Depressive episodes, characterized by sadness, anhedonia, and low energy and motivation, occur in both DD and BD and often complicate diagnostic decisions [5,6]. BD is uniquely characterized by episodes of hypomania or mania, marked by elevated or irritable mood, increased energy, and impulsivity [7,8]. Although not captured by traditional categorical approaches, subthreshold symptoms of depression and hypomania are common during and between episodes in both disorders [9–12]. Spectrum-based approaches to psychopathology offer a more nuanced understanding of mood dysregulation across diagnoses by capturing depressive and manic symptoms along a continuum [13–16].

Mood disorders are characterized by systematic biases in anticipating, encoding, and recognizing emotional stimuli [17–21]. Our previous work shows that anticipatory activation and connectivity during various tasks distinguish individuals with DD from those with BD [18,22]. Additionally, individuals with DD show a bias toward negative stimuli [23–25] and those with BD show a bias toward positive stimuli [26,27]. Recent research highlights that the processing of emotional faces in mood disorders is affected by top-down regulatory dysfunction [28] and depends on the valence of facial expressions [29] and mood symptoms [30]. For example, individuals with DD show aberrant responses in frontal, parietal, and subcortical regions to sad faces [31,32] and heightened amygdala responses during anticipation of aversive stimuli [33]. Those with depression and pronounced negative bias in future-oriented thinking showed higher frontopolar activation than their less pessimistic counterparts [34]. Individuals with BD show increased response in the striatal and prefrontal cortical [35,36] as well as visual regions [37] to emotional faces. Facial expressions provide rapid, socially salient cues important for social communication [38]. Therefore, linking depression and hypomania symptoms to emotional face anticipation and processing is key to understanding affective biases across the mood spectrum [17,39].

One underexplored factor that may influence stimulus anticipation and encoding is the vividness of visual imagery, the ability to form rich mental representations without external input [40]. Visual imagery recruits neural systems that substantially overlap with those involved in perception, including occipital, temporal, and parietal regions such as the angular gyrus [41–46]. It was shown that the vividness of visual imagery is related to task performance [47–50], which is possibly mediated by the frontopolar cortex and angular gyrus that support the integration of imagery into higher-order cognitive control and processing [51,52]. Depression is often associated with diminished vividness for positive imagery and greater vividness for negative imagery [53–55], whereas mania is associated with increased vividness of positive imagery [56,57]. Therefore, examining the relationship between vividness of visual imagery and brain activation during emotional stimulus anticipation and encoding across the spectrum of depressive and manic symptoms may provide valuable insights into the pathophysiology of mood disorders.

Building on prior work, we hypothesized that depressive spectrum symptoms would be associated with altered activation in frontopolar and parietal regions during anticipation and encoding of sad relative to happy faces, reflecting negative affective biases. Conversely, we expected that hypomania spectrum symptoms would predict heightened activation in the occipital cortex during encoding of happy relative to sad faces, consistent with enhanced responsiveness to positive emotional stimuli. We further predicted that these effects would be moderated by individual differences in the vividness of visual imagery, such that reduced vividness would amplify negative biases in depression, while heightened vividness would enhance positive biases in hypomania.

## 2 METHOD AND MATERIALS

### 2.1 Participants

The study was approved by the University of Pittsburgh Institutional Review Board (IRB number: STUDY20060265). Participants were recruited from the community, local universities, and medical centers. They provided written informed consent to participate in the study. Participants were right-handed, fluent in English, and sex-matched across groups. HC had no personal or family history of psychiatric disorders. Symptomatic individuals met DSM-5 criteria for depressive disorders (DD), such as major depressive or persistent depressive disorders, or bipolar disorder (BD) type I or II. We scanned a total of 170 participants (43 BD, 68 DD, and 59 HC). Of them, 2 BD, 5 HC, and 8 DD were removed from the analyses due to changing their diagnostic group (n=3), excessive motion (n=8), poor image quality (n=1), or did not respond to over 20% of trials during encoding (n=3), thus leaving 155 participants (41 BD, 60 DD, and 54 HC) in the analyses.

### 2.2. Clinical assessment

All diagnoses were made by a trained clinician and confirmed by a psychiatrist according to DSM-5 criteria using SCID-5 [58]. We assessed current depression symptoms (Hamilton Depression Rating Scale; HDRS-25)[59], current mania symptoms (Young Mania Rating Scale; YMRS) [60], and lifetime dimensional symptoms of depression and mania (Moods Spectrum self-report questionnaire; MOODS-SR) [9]. A total psychotropic medication load was calculated for each participant [22,61]. Exclusion criteria included history of head trauma with loss of consciousness, history of neurological, neurodevelopmental, systemic, or borderline personality disorders, meeting criteria for psychotic-spectrum disorders, substance use disorder in the past 6 months or current use of illicit substances determined by SCID-5-RV, and pre-scan saliva alcohol and urine drug screens, premorbid IQ<85 per North American National Adult Reading Test (NART) [62], standard MRI exclusion criteria (e.g., claustrophobia, cardiac or neural pacemakers, surgically implanted metal devices, metal braces, or other metal objects in their body), current pregnancy (either self-reported or positive pregnancy test at scan visit), inability to understand or speak English, left or mixed handedness (Annett’s criteria) [63] to ensure a uniform hemispheric dominance for interpretation of neuroimaging data, and the Young Mania Rating Scale (YMRS) [60] score > 8. Individuals with BD were excluded if they were in a manic state on the day of the scan.

### 2.3 Behavioral assessments

Participants performed a Cued Encoding of Emotional Faces task inside the scanner (Figure 1). The task consisted of 48 ten-second trials presented over 2 runs. Each trial started with the 4-second *anticipation* phase in which a concave line predicting happy faces, a convex line predicting sad faces, and a straight line predicting neutral faces. During this phase, participants were instructed to mentally prepare to encode and memorize a face with the cued emotion. Following the cue and a fixation cross presented for a variable inter-stimulus interval ranging from 1 to 2 sec, a face stimulus was presented for up to 1.5 sec. The face pictures were taken from the NimStim [64] and KDEF [65] databases. Participants were instructed to rate the image as pleasant or unpleasant as quickly as possible and memorize the faces for the subsequent memory test. The responses were made by pressing a corresponding button with the index finger on one hand for pleasant images and on the other hand for unpleasant images. The hand assignment for pleasantness rating was counterbalanced across subjects. Participants were informed that there was no right or wrong answer. The rest between the trials ranged from 3290 to 4783 msec. During practice trials, participants learned the association between abstract shapes and facial emotional expressions.

**Figure 1.**
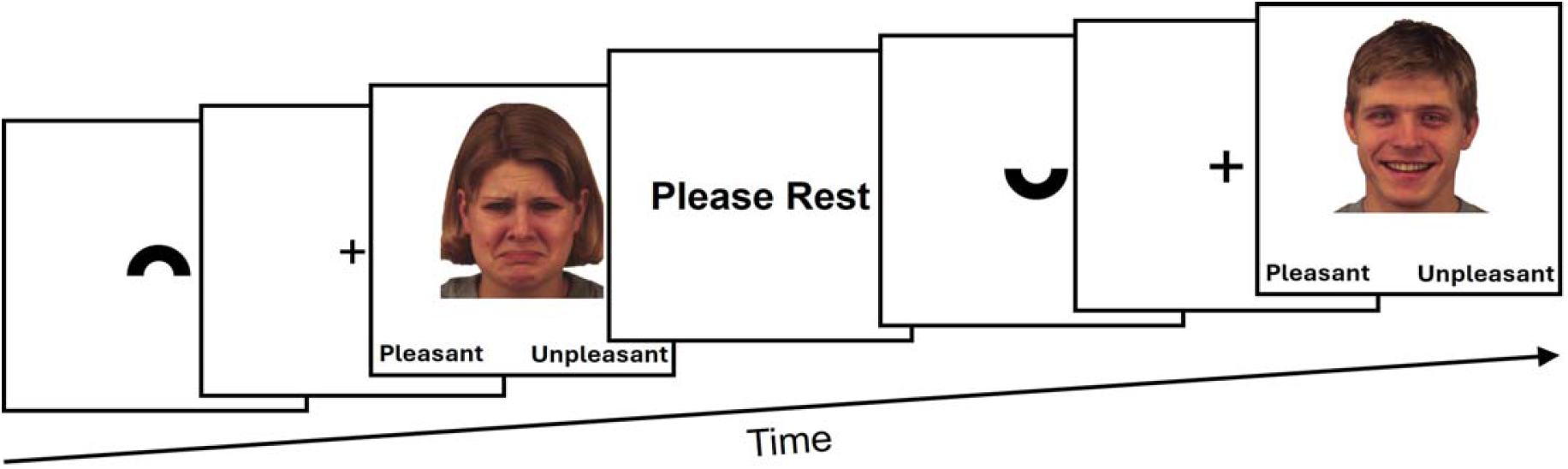
Experimental paradigm. The images BF26SAS and BM23HAS were taken from the KDEF[65].

### 2.4 Neuroimaging data acquisition

The neuroimaging data were collected at the University of Pittsburgh/UPMC Magnetic Resonance Research Center using a 3T Siemens Prisma scanner with a 64-channel head coil and named according to the ReproIn convention[66]. The EPI data were collected in the anterior-to-posterior direction using a multi-band sequence (factor=8, TR=800ms, resolution=2×2×2mm, FOV=210, TE=30ms, flip angle=52°, 72 slices, 315 volumes). High-resolution T1w images were collected using the MPRAGE sequence (TR=2400ms, resolution=0.8×0.8×0.8mm, 208 slices, FOV=256, TE=2.22ms, flip angle=8°). Field maps were collected in the AP and PA directions using the spin echo sequence (TR=8000, resolution=2×2×2mm, FOV=210, TE=66ms, flip angle=90°, 72 slices).

### 2.5 Data analyses

#### 2.5.1 Clinical data analysis

Demographic and clinical characteristics were compared between groups using t- and chi-square tests. A total psychotropic medication load was calculated for each participant [22,61].

#### 2.5.2 Behavioral data analysis

All repeated measures analyses were conducted using a mixed effects model using the ‘*lme4*’ package in R [67] with p-values and summary tables calculated using the ‘*lmerTest’* package in R [68]. In all models, participants were treated as a random factor, and their age, sex, and IQ were used as covariates. For significant effects, the contrasts and means were estimated from the mixed effects models using the ‘*modelbased’* package in R [69].

#### 2.5.3 Neuroimaging data analysis

##### Preprocessing

The DICOM images were converted to a BIDS[70] dataset using heudiconv version 0.5.4[71]. Data quality was examined using mriqc 0.15.1[72]. The data were preprocessed using fmriprep 20.1.1 [73]. Specifically, T1w images were skull-stripped, and brain surfaces were reconstructed using recon-all (FreeSurfer 6.0.1) [74], and brain masks were generated. Preprocessing steps included generating a reference volume and its skull-stripped version using a custom methodology of fmriprep, estimating head-motion parameters with respect to the BOLD reference before any spatiotemporal filtering using mcflirt [75], and applying slice-time correction using 3dTshift [76], (AFNI 20160207; RRID:SCR_005927). Fieldmaps were estimated with 3dQwarp [76] (AFNI 20160207) based on two spin echo images collected with opposing phase-encoding directions (i.e., anterior-to-posterior, and posterior-to-anterior). Based on estimated susceptibility distortion, a corrected EPI (echo-planar imaging) reference was calculated for more accurate co-registration with the anatomical reference. The BOLD reference was co-registered to the T1w reference using bbregister (FreeSurfer) [74] (RRID:SCR_001847) which implements boundary-based registration [77]. Co-registration was configured with six degrees of freedom. The BOLD time-series were resampled onto the fsaverage surfaces (FreeSurfer reconstruction nomenclature) and onto their native space by applying a single, composite transform to correct for head-motion and susceptibility distortions. The BOLD time-series were resampled into standard space, generating a preprocessed BOLD image in MNI152NLin2009cAsym space. Automatic removal of motion artifacts using independent component analysis (ICA-AROMA) [78] was performed on the preprocessed BOLD after removal of non-steady state volumes and spatial smoothing with an isotropic, Gaussian kernel of 6mm FWHM (full-width half-maximum). After that, global signals within the CSF and WM were extracted and regressed out from preprocessed BOLD data, and high-pass temporal filter (90-sec cutoff) were applied. All resamplings were performed with a single interpolation step by composing all the pertinent transformations (i.e., head-motion transform matrices, susceptibility distortion correction when available, and co-registrations to anatomical and output spaces). Gridded (volumetric) resamplings were performed using antsApplyTransforms (ANTs), configured with Lanczos interpolation to minimize the smoothing effects of other kernels [79].

##### Subject-level analysis

Subject-level statistical maps were computed using FSL 6.0.3 installed system-wide on the workstation with GNU/Linux Debian 10 operating system with NeuroDebian repository [80]. A hemodynamic response was modeled using a gamma density function. A subject-level model included 6 explanatory variables that included happy, sad, and neutral faces and corresponding cues. The contrasts of interest included happy vs. sad stimuli during anticipation and encoding phases of the task. Positive differences (increases) show greater activation for happy vs. sad (happy>sad), while negative differences (decreases) show lower activation for happy vs. sad (happy<sad).

##### Group-level analyses

The first analysis conducted across all 155 participants calculated the activation maps for all anticipatory cues relative to resting baseline and for all face stimuli relative to resting baseline. The images of the four resulting maps (activation and deactivation during anticipation and stimulus encoding) were obtained using the *swe* (Sandwich Estimator) approach [81] using nonparametric permutation inference with 5000 permutations, Threshold-Free Cluster Enhancement (TFCE) correction, and the FWE-corrected p-values threshold setup to p=0.01. After that, the brain images of anticipatory processing were overlayed with the brain images of stimulus encoding to produce four masks representing regions that activated during both anticipation and encoding, deactivated during both anticipation and encoding, activated during anticipation but deactivated during encoding, and deactivated during anticipation but activated during encoding.

The spectrum depression-by-VVIQ and the spectrum hypomania-by-VVIQ interactions and main effects in the four masks of interest were examined for the happy vs. sad contrast during anticipation and encoding phases of the task. The swe approach[81] using nonparametric permutation inference with 5000 permutations, Threshold-Free Cluster Enhancement (TFCE) correction, and the FWE-corrected p-values threshold setup to p=0.05 was used. Age, gender, and IQ were used as covariates to account for potential individual differences associated with these variables. Functional localization was determined using the Harvard-Oxford cortical and subcortical structural atlases and visualized using *fslviewer and mricron* [82]. The mean percent signal changes were extracted from the clusters of voxels showing a significant interaction or main effects and used in the follow-up analyses using a *featquery* tool in FSL.

## 3 RESULTS

### 3.1 Clinical and behavioral

The study sample is described in Table 1. The groups did not differ in sex composition. The mean age of participants was under 30 years, but participants in the BD group were younger than HC. Mean IQ exceeded 100 across all groups, although participants with BD had higher scores than HC. The BD and DD groups did not differ in age of illness onset, illness duration, number of individuals with comorbid psychiatric disorders, YMRS, or lifetime spectrum depression scores. Individuals with BD had significantly more mood episodes, more severe lifetime spectral symptoms of hypomania, and had a higher medication load (medload) than those with DD (*p* < 0.05 for all comparisons).

**Table 1.** Demographic and clinical characteristics.

|  | BD | DD | HC | Statistics |
| --- | --- | --- | --- | --- |
| Baseline visit |  |  |  |  |
| N | 41 | 60 | 54 |  |
| Age at scan (years) | 25.5(4.3) | 27.8(6.6) | 28.8(6.6) | F(2,152)=3.5, $p=0.03$<br>HC vs. BD: $t(93)=2.8$, $p=0.007$ |
| Sex (number of female) | 35 | 46 | 41 | n.s. |
| IQ | 110.96(5.8) | 109.6(7.1) | 107.4(5.7) | F(2,152)=4.42, $p=0.014$<br>HC vs. BD: $t(93)=-3.12$, $p=0.002$ |
| HRSD25 | 9.9(7.6) | 13.2(7.2) | 1.57(1.8) | BD vs. DD: $t(99)=-2.21$, $p=0.03$ |
| YMRS | 1.46(2.09) | 1.2(1.5) | 0.22(0.7) | BD vs. DD: n.s. |
| MOODs-SR depression lifetime | 19.9(4.1) | 18.5(4.06) | 2.1(2.4) | BD vs. DD: n.s. |
| MOODs-SR mania lifetime | 17.9(4.9) | 9.3(5.8) | 3.3(2.8) | BD vs. DD: $t(99)=7.8$, $p<0.001$ |
| Illness onset (age in years) | 14.8(3.7) | 15.13(4.7) | na | BD vs. DD: n.s |
| Illness duration (years) | 10.7(5.14) | 12.63(7.06) | na | BD vs. DD: n.s |
| Total number of mood episodes | 11.3(8.97) | 6.7(13.66) | na | $t(98)=4.04$, $p<0.001$ |
| Number of individuals with comorbid psychiatric disorders | 22 | 38 | na | n.s. |
| Number of individuals taking psychotropic medications | 36 | 40 | na | $\chi^2(1)=4.76$, $p=0.03$ |
| Medload in individuals taking psychotropic medications | 3.28(1.56) | 2.25(1.26) | na | BD vs. DD: $t(74)=3.18$, $p=0.002$ |
| VVIQ (higher score reflects lower vividness of visual imagery) | 35.66(13.38) | 33.85(13.49) | 33.13(12.58) | n.s. |

*Note:* The table reports the means and standard deviations (SD) that are reported in parentheses. Clinical variables measuring depression and mania symptoms were compared between BD and DD groups only due to consistently low scores in HC.

Lifetime hypomania spectrum symptoms, but not depression spectrum symptoms, predicted VVIQ scores after the diagnostic group, age, sex, and IQ were accounted for (F(1,145)=4.9, p=0.03, t= −2.28, p= 0.024). More symptomatic individuals across diagnoses had lower VVIQ scores that corresponded to higher vividness of visual imagery (Figure 2). Using the *boxplot*.*stats* function in R, we identified 4 outliers with high VVIQ scores (VVIQ score>67, 1 HC, 0 BD, and 3 DD). The negative slope persisted even after removing these outliers from the model (F(1,141)=4.13, p=0.044, t= −2.07, p= 0.04).

**Figure 2.**
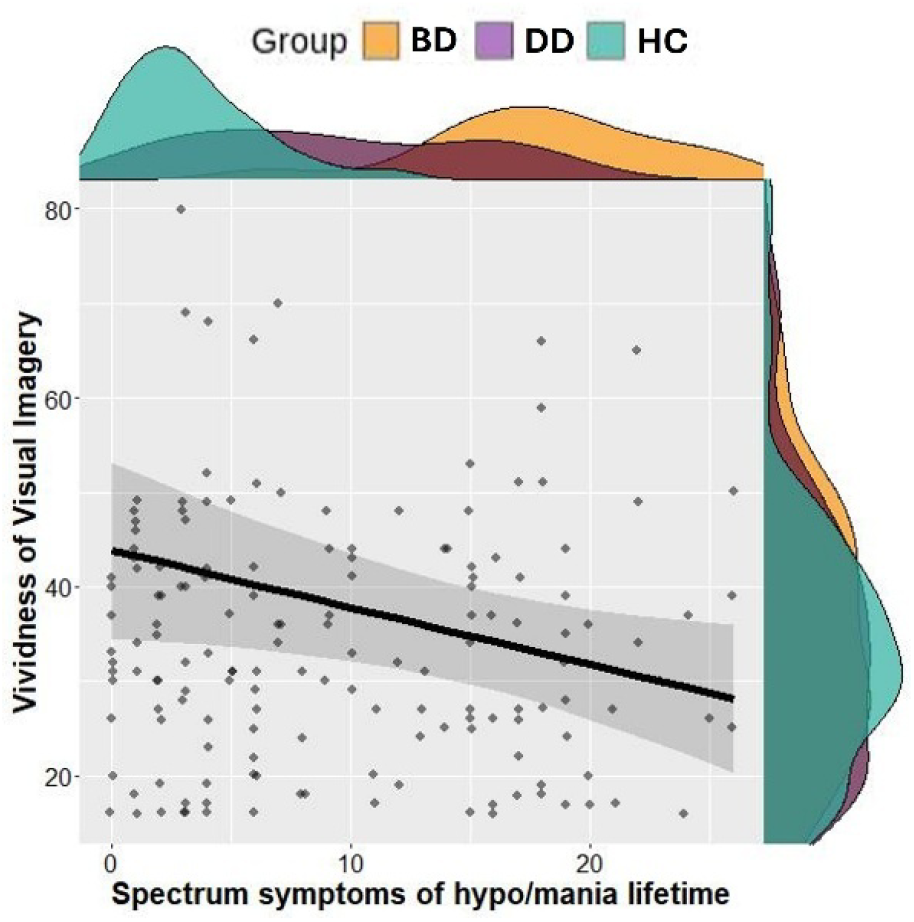
The relationship between lifetime spectrum hypomania symptoms and VVIQ score (higher score indicates low vividness of imagery).

The mixed effects model examining lifetime spectrum hypo/mania symptoms, vividness of visual imagery and valence (happy and sad) of facial emotional expression revealed a significant 3-way interaction (*F*(1,4728)=5.5, *p*=0.02), a significant spectrum mania and emotion interaction (*F*(1,4728)=9.0, *p*=0.003), and the main effect of facial emotions (*F*(1,4728)=11.1, *p*<0.001) with slower responses to the sad faces. While RT for happy faces was slower for individuals with higher spectrum hypomania independently of VVIQ scores, RT for sad faces was slower for those with high levels of spectrum hypomania and lower vividness of imagery (higher VVIQ scores), and for those with lower levels of spectrum hypomania and higher vividness of imagery (lower VVIQ scores). The mixed effects model examining lifetime spectrum depression symptoms, vividness of visual imagery and valence (happy and sad) of facial emotional expression revealed a significant main effect of facial emotions (*F*(1,4728)=4.6, *p*=0.03) with slower responses to sad faces, but no interaction effects. These results are depicted in Figure 3.

**Figure 3.**
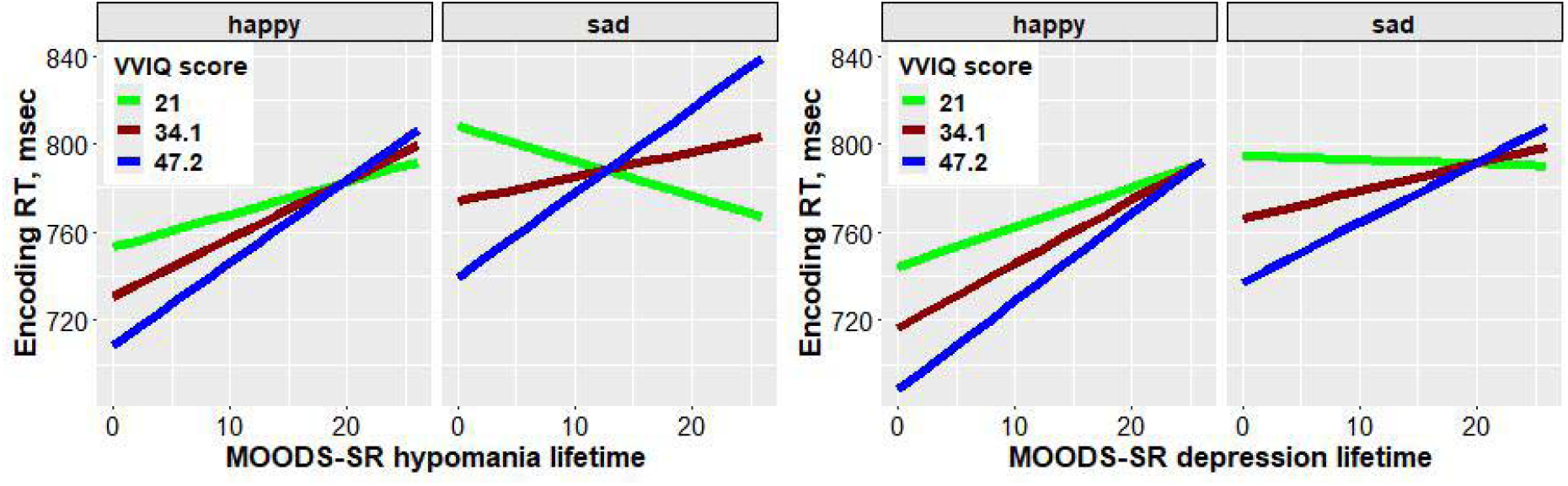
The relationship between spectrum mood symptoms, VVIQ scores and stimuli valence on encoding RT.

### 3.2 Neuroimaging

We identified four masks of interest: brain areas where both cues and faces elicited activation increases, areas where both cues and faces elicited activation decreases, areas with activation increases for cues but decreases for faces, and areas with activation decreases for cues and increases for faces (Figure 4).

**Figure 4.**
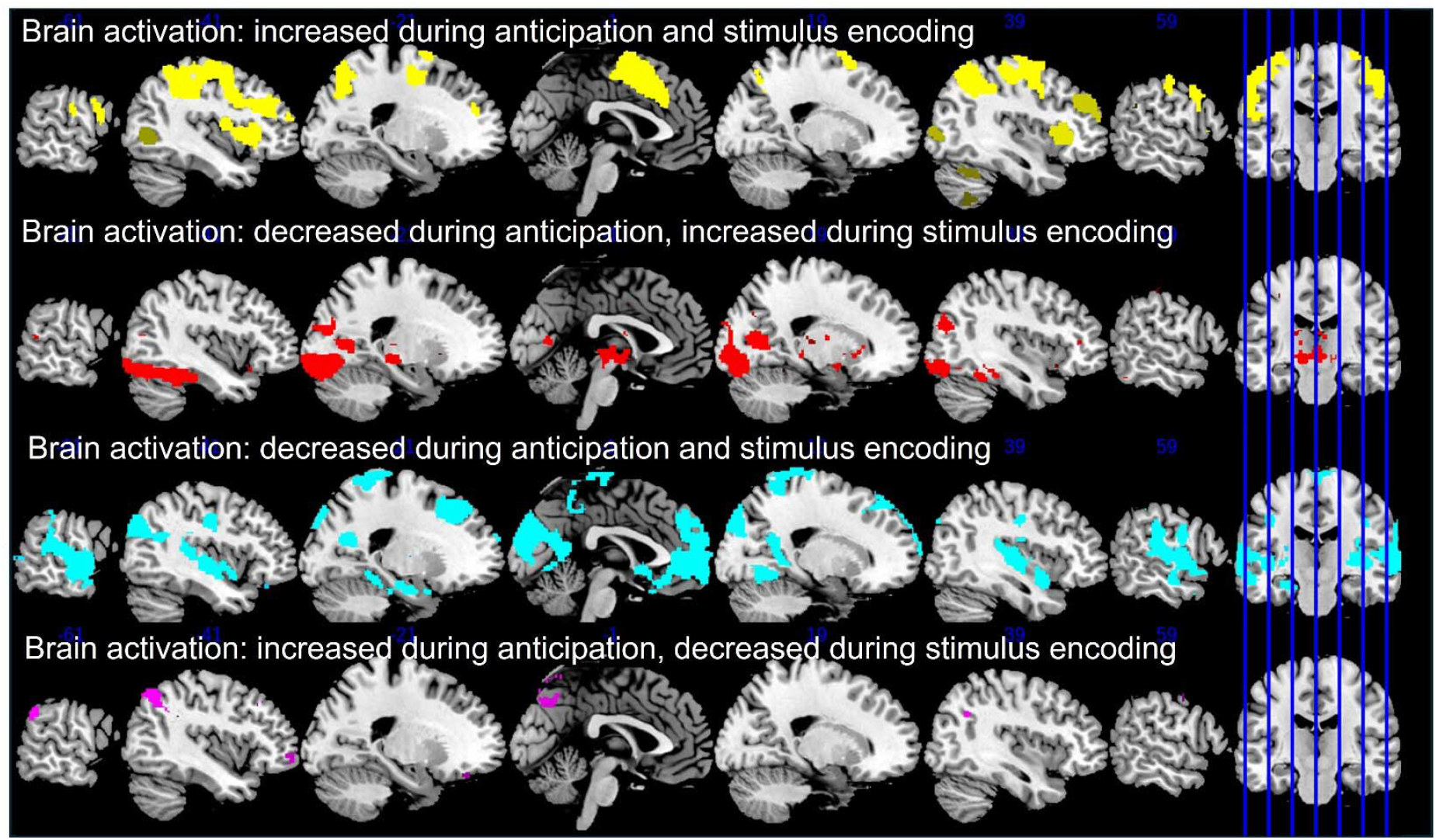
Brain activation patterns during stimulus anticipation and encoding activation. The picture on the right with vertical lines shows the location of each slice on the brain.

#### Task-positive networks

The mask that consisted of regions showing activation increases during both anticipation and encoding (illustrated in yellow on Figure 4) included prefrontal cortex, precentral and postcentral gyri, insular cortex, anterior cingulate and paracingulate cortices, superior parietal lobule, lateral occipital cortex, and cerebellum.

The mask that consisted of regions showing activation increases during encoding but decreases during anticipation included temporo-occipital regions (inferior temporal gyrus, temporal and tempo-occipital fusiform cortices, occipital fusiform and lingual gyri, intracalcarine and lateral occipital cortices, occipital pole), and subcortical regions (hippocampus, amygdala, thalamus, putamen, and caudate) (illustrated in red on Figure 4).

#### Task-negative networks

The mask that consisted of regions showing activation decreases during both stimulus anticipation and encoding (illustrated in blue on Figure 4) included following regions: frontal pole, medial aspects of superior/ middle frontal and temporal gyri, postcentral, supramarginal, superior parietal, lateral occipital cortex, subcallosal, paracingulate gyrus, anterior and posterior cingulate, cuneus and precuneus, lingual gyrus, central and parietal opercular, planum polare, Heschl’s gyrus, planum temporale, supracalcarine, occipital pole, as well as putamen and hippocampus.

The mask that consisted of regions showing activation decreases during encoding but increases during anticipation included frontal pole, prefrontal cortex, supramarginal and angular gyri, and lateral occipital cortex (illustrated in magenta on Figure 4).

#### 3.2.1 Spectrum depression

We found a significant spectrum depression by VVIQ score interaction effect on the differences in brain activation during anticipation of happy vs. sad faces in the left frontal pole region (nvox=67, [-2, 58, 2], t-max= 5.38) located in the mask combining anticipation and encoding related brain deactivation. Individuals with higher spectrum depression and higher VVIQ scores (lower vividness of imagery) showed greater activation during anticipation of happy than sad faces, while those with lower depression scores and higher VVIQ scores showed greater activation during anticipation of sad than happy faces. The depression by VVIQ interaction effect was also found during face encoding in the left angular gyrus (nvox=24, [-50, −58, 46], t-max=4.5) located in the cue increase-encoding decreases mask (Figure 5).

**Figure 5.**
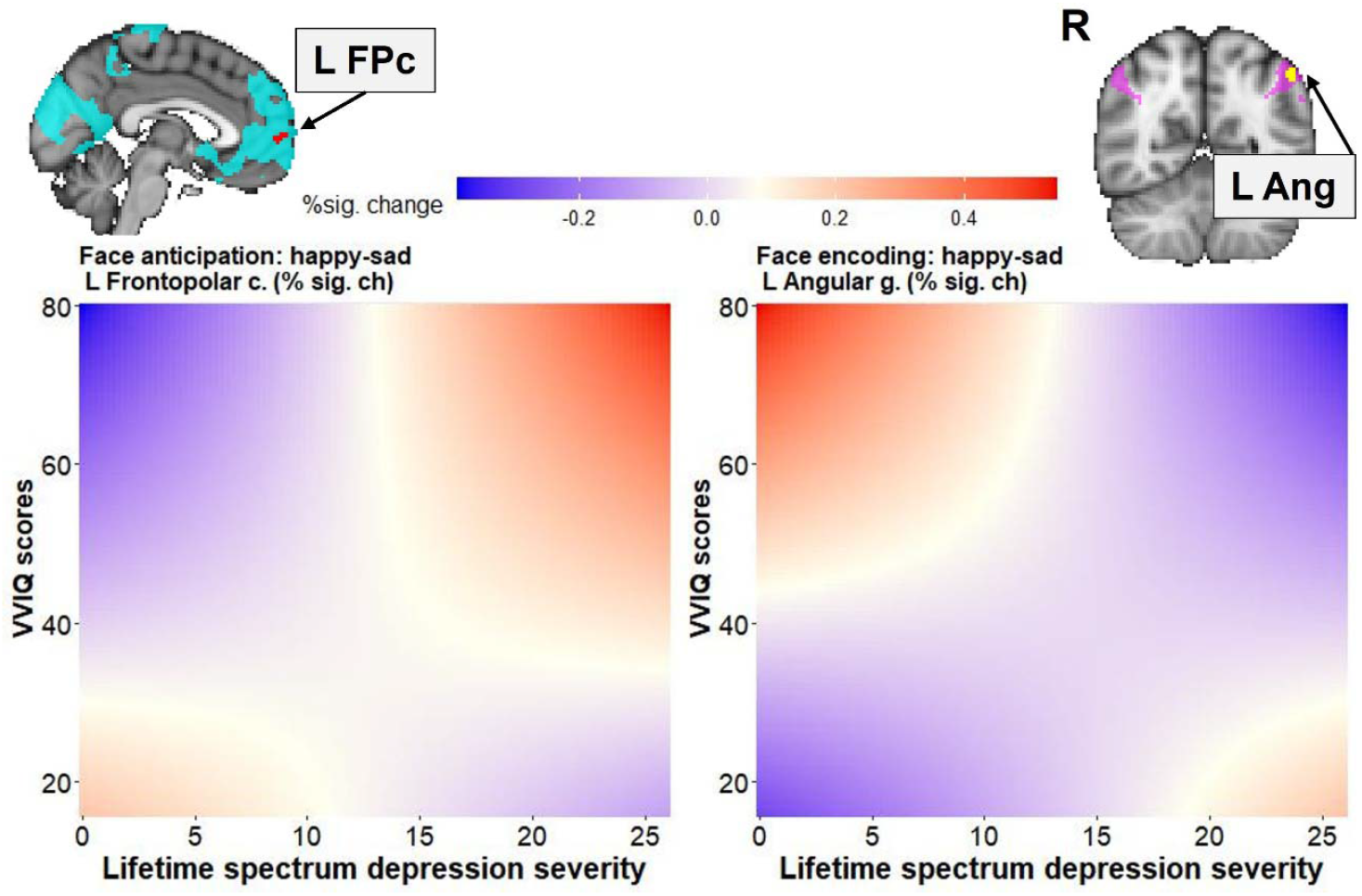
Spectrum depression by VVIQ score interaction effects on happy minus sad activation differences during face anticipation and encoding. The red color of the heat map illustrates happy>sad, while the blue color illustrates happy<sad differences. The blue brain mask represents the regions of activation decrease during both face anticipation and processing, with red representing the area of significant activation in the left frontopolar cortex (L FPc). The magenta brain mask represents the regions activating during face anticipation but deactivating during face encoding, with yellow representing the area of significant activation in the left angular gyrus (L Ang). “R” stands for the right hemisphere.

#### 3.2.2 Spectrum mania

There was no significant spectrum hypomania by VVIQ score interaction effect. The main effect of hypomania was observed in the R precentral/postcentral gyrus (nvox=18, [44, −16, 52], t-max= 4.74) that was a part of the mask that included anticipation and encoding-related increases in brain activation. Higher spectrum hypomania scores were associated with greater brain activation during anticipation of sad vs. happy faces. The positive relationship between spectrum hypomania and happy vs. sad differences was observed in the cue increase - encoding decrease mask in the superior division of the left LOC (nvox=71, [-40, −62, 46], t-max= 3.89): individuals with high spectrum hypomania scores showed greater activation during encoding of happy vs. sad faces (Figure 6).

**Figure 6.**
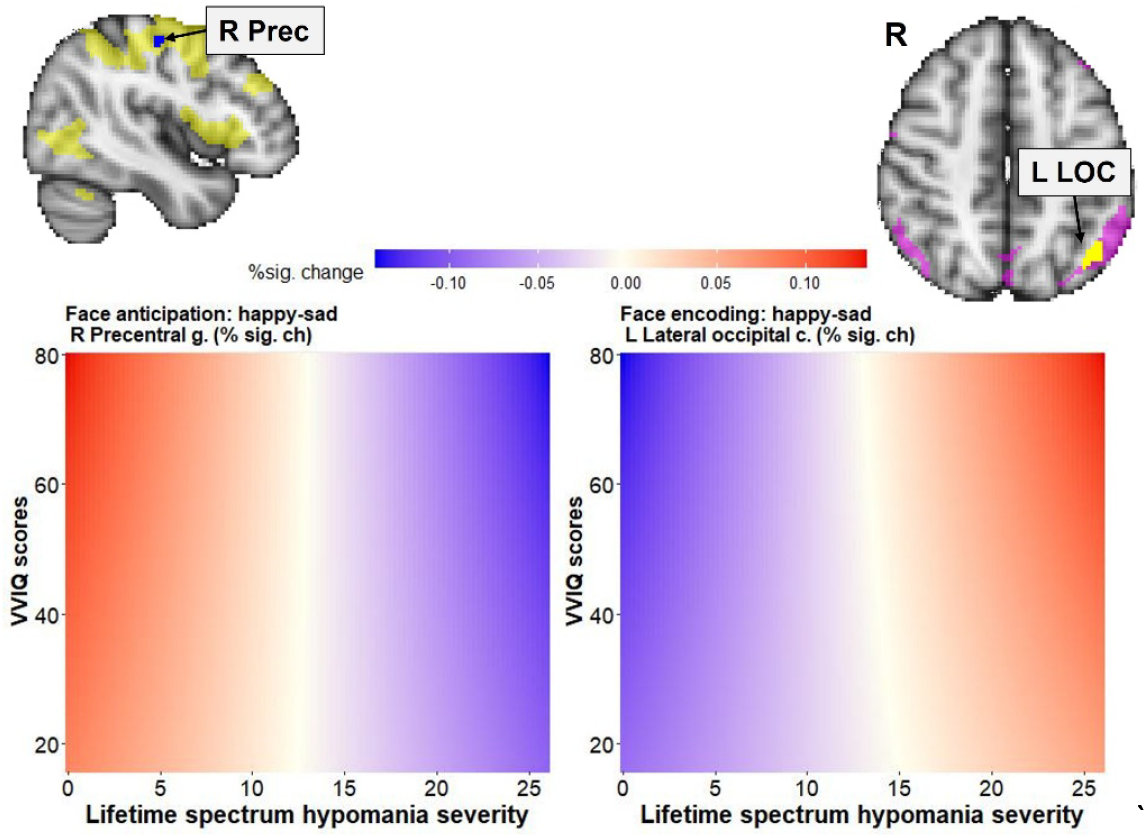
The effect of spectrum mania on happy vs. sad activation differences during face anticipation and encoding. The red color of the heat map illustrates happy>sad, while the blue illustrates happy<sad activation. The yellow brain mask represents the regions with activation increases during both face anticipation and encoding, with blue representing the area of significant activation in the right precentral gyrus (R Prec). The magenta brain mask represents the regions activating during face anticipation but deactivating during face encoding, with yellow representing the area of significant activation in the left lateral occipital cortex, superior division (L LOC). “R” stands for the right hemisphere.

A positive relationship between spectrum hypomania and happy vs. sad face activation difference was observed in the combined decreases mask in the occipital cortex (left occipital pole: nvox=49, [-10, −92, 22], t-max= 3.7; right cuneus/precuneus: nvox=31, [0, −80, 40], t-max= 3.46; right intracalcarine cortex: nvox=113, [6, −88, 12], t-max= 4.8) (Figure 7). Individuals with higher hypomania scores across diagnoses showed greater activation during encoding of happy vs. sad faces, while those with lower spectrum hypomania showed greater activation for sad vs. happy faces.

**Figure 7.**
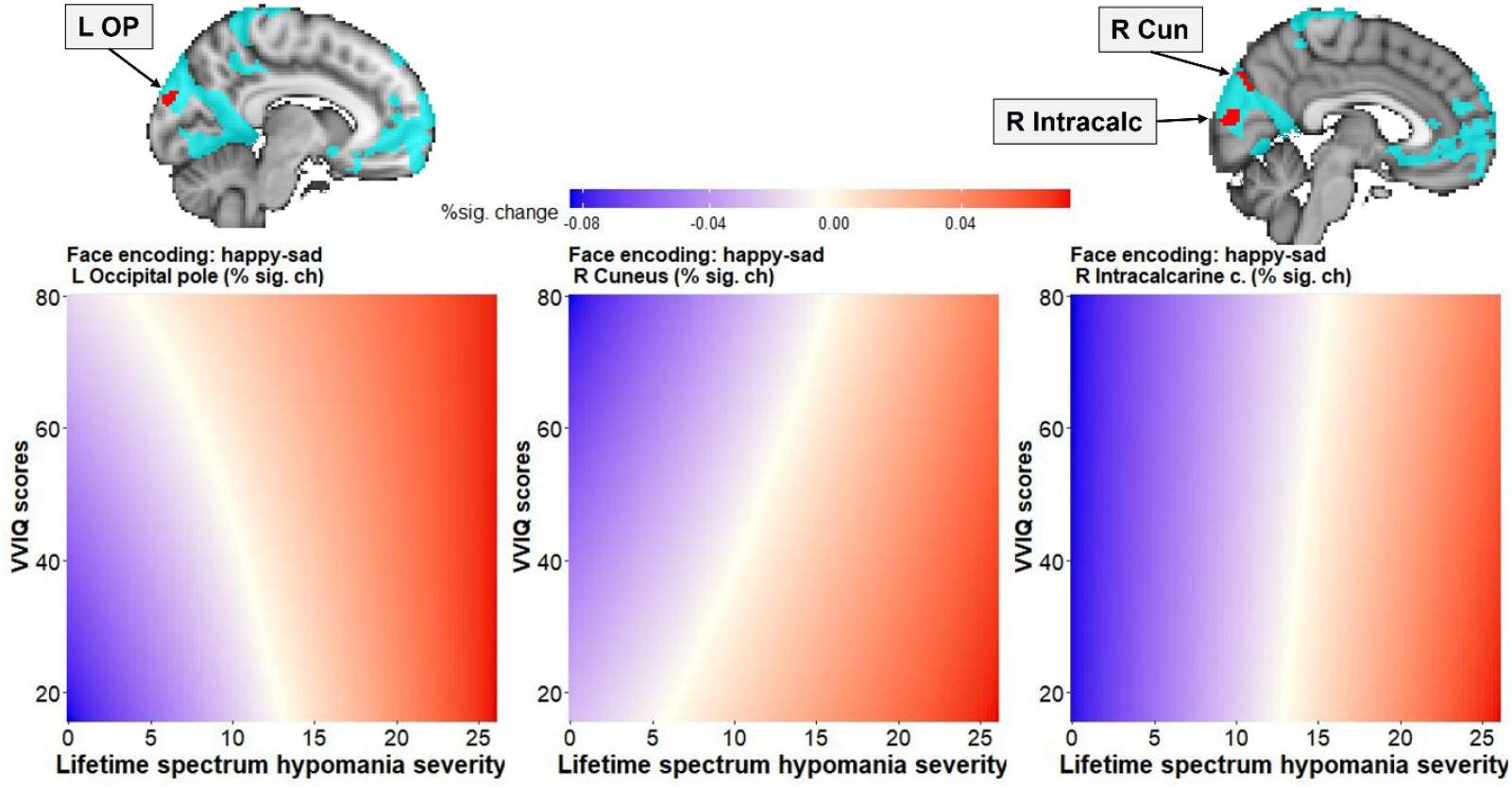
The effect of spectrum mania on happy vs. sad activation differences during face encoding. The red color of the heat map illustrates happy>sad, while the blue illustrates happy<sad activation. The blue brain mask represents the regions decreasing activation during both face anticipation and encoding, with red representing the areas of significant activation in the left occipital pole (L OP), right cuneus (R Cun), and right intracalcarine (R Intracalc) cortices. “R” stands for the right hemisphere.

#### 3.2.3 Exploratory analysis

The difference in RT for making pleasant/unpleasant judgments for happy vs. sad faces did not correlate with the happy-sad differences in brain activation in the brain regions reported above.

## 4 DISCUSSION

This study examined how individual differences in vividness of visual imagery moderate the relationship between lifetime spectrum symptoms of depression and hypomania and neural responses during anticipation and encoding of happy and sad faces in individuals with mood disorders and healthy controls. Consistent with previous work, we found that greater lifetime hypomania was associated with higher imagery vividness [83–86]. While imagery vividness and spectrum depression were unrelated to RT during stimulus encoding, the interaction between these two variables was associated with activation in the left frontopolar cortex during anticipation and the left angular gyrus during encoding of happy vs. sad faces. In contrast, RT during stimulus encoding was associated with the interaction between spectrum hypomania, imagery vividness, and stimulus valence. Slower responses to happy faces were observed in individuals with higher spectrum hypomania independently of imagery vividness. Slower responses to sad faces were observed in individuals with higher hypomania and lower imagery vividness, and those with lower hypomania but higher imagery vividness.

These findings were not replicated at the neural level, though. Spectrum hypomania, but not imagery vividness, was associated with the differences in brain activation for happy vs. sad faces observed in the right precentral gyrus during anticipation and in the left superior lateral occipital cortex, occipital pole, cuneus, and intracalcarine cortex during encoding. Overall, our findings are in line with dimensional models of mood psychopathology [10,15,16] and suggest that mood spectrum traits across diagnoses and visual imagery vividness bias anticipatory and subsequent encoding processes. Specifically, depressive traits were associated with engagement of frontopolar–parietal cortices, while hypomanic traits were associated with engagement of sensorimotor and occipital cortices. These findings also align with existing evidence demonstrating that depression is associated with a biased encoding towards negative information [87,88], whereas hypomania tends to amplify the encoding of positive information [83,85,86]. These patterns align with broader evidence that anticipatory activity biases subsequent encoding [89,90] and with accounts that imagery can support both anticipation and encoding by recruiting overlapping networks [40,42].

### Depression, imagery vividness, and anticipatory processing

The interaction effect between spectrum depression and vividness of visual imagery was observed in the left frontopolar cortex during stimulus anticipation and the left angular gyrus during stimulus encoding. Both regions are components of the default mode network [91] that deactivate during stimulus encoding and play a role in self-referential thought [92–94] and future thinking [95,96]. Interestingly, the left frontal pole showed deactivation during both anticipation and stimulus encoding, thus reflecting potential preparatory brain modulation [97]. The left angular gyrus deactivated during encoding, but activated during anticipation, which could be consistent with the account of resource allocation based on expected cognitive load [18,98].

The finding that anticipation-related activation in the left frontopolar cortex was greater for happy than sad stimuli in individuals with higher spectrum depression and lower imagery vividness suggests compensatory recruitment of this region when positive content mismatches lifetime experience with negative emotions, and the ability to generate vivid positive images is low. This is consistent with depressive expectancy biases and the need for greater top-down control to process positive input [87,99,100]. Conversely, individuals with low spectrum depression and low vividness showed greater activation in the frontopolar region during anticipation of sad vs. happy faces, suggesting that depressive traits can modulate the direction of frontopolar engagement while preparing to process emotional information.

In contrast, the left angular gyrus exhibited greater activation for sad vs. happy faces during stimulus encoding in individuals with higher spectrum depression and lower imagery vividness, but showed greater activation for happy vs. sad faces in individuals with low spectrum depression scores and low vividness. Diminished vividness in more depressed individuals may strengthen negative expectancies, whereas lower depressive symptoms permit preferential integration of positive cues even when imagery is weak. This dynamic reflects the brain’s complex mechanisms for predicting and responding to environmental stimuli and supports the idea that reduced imagery vividness interacts with depressive traits to bias anticipatory processes toward negative outcomes [20,21], ultimately affecting the encoding of social stimuli such as faces.

### Hypomania and positive encoding biases

Our behavioral findings demonstrated a greater reaction time difference between sad and happy faces (slower for sad) in participants with higher hypomania scores, thus suggesting that, for these individuals, encoding sad faces was more difficult than happy faces. A more challenging task typically requires greater activation within the task-positive network. During anticipation, higher lifetime spectrum hypomania was related to greater activation in the right precentral gyrus for the cues predicting sad vs. happy faces suggesting that individuals with higher spectrum hypomania pre-activated this region during anticipation of sad faces to support more efficient processing during subsequent encoding. The precentral gyrus is the region showing activation increases during both stimulus anticipation and encoding, and is involved in motor responses initiation, motor imagery, and response anticipation [101]. Our findings suggest that the readiness to respond could be biased toward sad faces, thus reflecting a potential conflict with lifetime experiences of spectrum hypomania. This activation pattern aligns with our previous findings and those of others that preparatory activation establishes neural states that facilitate task execution [33,98,102–105].

During encoding, hypomanic spectrum symptoms correlated with greater activation for happy vs. sad faces in the occipital regions that belong to a task-negative network that deactivates during task performance. Occipital and intracalcarine cortices have been implicated in conscious perception [46]. Increased recruitment of these areas may reflect enhanced perceptual and mnemonic representation of happy faces. These results suggest that spectrum hypomania traits enhance positive-valence encoding via reduced deactivation of visual cortices, consistent with reports of positivity-biased responses [36,106] and reduced sensitivity to negative social cues [57,107] observed in BD. The dimensional association between hypomanic traits and occipital recruitment is also consistent with evidence that individuals at risk for BD exhibit heightened reward sensitivity and positive affective biases even outside of acute episodes [108,109].

### Implications for imagery, memory, and mood spectrum models

Our findings underscore the importance of considering visual imagery as a modulatory factor in affective processing. Prior research shows that imagery vividness enhances working and episodic memory [47,48,50], and amplifies emotional experiences [57]. In our study, vividness of visual imagery interacted with depressive but not hypomanic traits, suggesting distinct pathways through which imagery influences negative versus positive biases.

Clinically, these results suggest that imagery-based interventions [57,110] could benefit individuals with depressive traits, by enhancing anticipatory simulation of positive outcomes and thereby correcting negative biases in encoding. In bipolar spectrum populations, interventions may need to address the opposite imbalance, mitigating excessive positive encoding biases that could contribute to risk-taking and social misjudgments. The frontopolar, angular gyrus, and visual cortices may be potential treatment targets for neuromodulation interventions to reduce depression and hypomania symptom severity.

## Limitations and future directions

This study has several limitations that should be considered. First, our sample combined HC and individuals with DD and BD, consistent with spectrum-based approaches. Future work should test whether these effects generalize across clinical subtypes and mood states [28,30]. Second, imagery vividness was self-reported using the VVIQ. Including objective imagery tasks would provide a more precise measure of imagery strength and vividness, thus reducing potential reporting biases. Prospective studies should manipulate imagery vividness (e.g., via training or by using online imagery prompts during cueing) to test causal effects on anticipatory and encoding processes. Third, inconsistent with our expectations, we found no interaction between imagery vividness and spectrum hypomania symptoms. One explanation for the lack of interaction could be collinearity between spectrum hypomania and imagery vividness due to their intercorrelation. Future studies should investigate this relationship further, potentially employing designs that disentangle their common and unique contributions to the processing of emotional stimuli. Finally, this paper focuses on anticipation and encoding of happy and sad faces without considering subsequent memory performance. Findings on vividness of visual imagery and memory for emotional faces will be reported separately.

## Conclusion

In summary, our study links lifetime spectrum symptoms to brain responses to emotional cues and faces, reflecting trait-like neural differences associated with mood-spectrum experiences. The study provides novel evidence that lifetime depressive and hypomanic spectrum symptoms across individuals with and without mood disorders differentially interact with visual imagery vividness to modulate anticipatory and encoding processes of emotional faces. Depressive traits coupled with low imagery vividness bias anticipatory processing toward negative outcomes, while hypomanic traits enhance positive encoding biases in visual cortices. These results highlight the utility of dimensional, spectrum-based approaches to mood psychopathology and underscore imagery vividness as a promising transdiagnostic target for interventions in mood disorders.

## FUNDING ACKNOWLEDGEMENTS

This work was supported by grants from the National Institute of Health R01MH114870 to A.M.

## ACKNOWLEDGMENTS

The authors thank participants for taking part in this research study.

## AUTHOR STATEMENT

A.M. – Conceptualization, Data curation, Formal analysis, Funding acquisition, Investigation, Project administration, Supervision, Validation, Visualization, Writing – original draft, Writing – review & editing

S.S. and R.M. – Data curation, Investigation, Project administration, Validation, Writing – review & editing

S.I. – Conceptualization, Methodology, Validation, Writing – review & editing

H.A.S. – Conceptualization, Investigation, Supervision, Validation, Writing – review & editing.

All authors have read and approved the final version of the manuscript and agreed to be accountable for all aspects of this work.

## DATA AVAILABILITY

The data that support the findings of this study are openly available in The National Institute of Mental Health Data Archive (NDA) at https://nda.nih.gov/, reference number Collection ID: 2908.

